# Prediction of In-Hospital Mortality for ICU Patients with Heart Failure

**DOI:** 10.1101/2024.06.25.24309448

**Authors:** J. Zhang, H. Li, N. Ashrafi, Z. Yu, G. Placencia, M. Pishgar

## Abstract

Heart failure affects millions of people worldwide. It greatly reduces quality of life and is associated with high mortality rates. Despite extensive research, the statistical connection between heart failure and mortality rates for ICU patients remains underexplored, indicating the need for improved prediction models.

This study identified 1,177 patients over 18 years old from the MIMIC-III database using ICD-9 codes. Preprocessing consisted of handling missing data, deleting duplicates, treating skewness, and oversampling to alleviate data imbalances. 18 features were selected within a LightGBM model by checking Variance Inflation Factor (VIF) values, LASSO Regression, and univariate analysis. The final output of the LASSO Logistic Regression model had the highest test AUC-ROC of 0.8766 (95% CI 0.8065 - 0.9429) and accuracy of 0.7291 compared to other baseline models, including Logistic Regression, Random Forest, LightGBM, Support Vector Machine (SVM), and Decision Trees. All models demonstrated good calibration with relatively low Brier scores, highlighting their reliability in predicting in-hospital mortality.

Our models predicted deaths of heart failure ICU patients better than the best results found in both literature and baseline models. These results were based on preprocessing missing values via improved imputation strategies and improved feature selection based on an expanded literature search and improved experiences selecting key features. With the Grid-Search, we had a near-perfect predictive model. These methods greatly increased the predictive accuracy of in-hospital mortality in ICU patients with heart failure.

## 1 INTRODUCTION

Heart failure (HF) affects approximately 6.5 million Americans aged 20 years and over making it a critical field of study. Those with HF experience severe symptoms including difficulty breathing, excessive coughing, and ultimately early death in about a quarter of cases at 1 year. [1] Hospitalization of HF patients often involve a serious infection, e.g. sepsis, in 20% of patients admitted to the ICU with life-threatening conditions. [2] Without further treatment, in-hospital mortality of HF patients will continue to be almost 10% based on [3].

Predictive models forecasting HF patient death in ICUs are critical. The introduction of Electronic Health Records (EHR) has positively affected patients’ treatment by utilizing information generated by the application of the data, improving performance, and increasing efficiency. [4, 5] Machine learning (ML) methods can find patterns and correlations among various features in large, complex data sets. This has improved doctors’ ability to diagnose and cure heart failure. [6 - 8] Several studies attempted to develop models to forecast HF patient deaths in ICUs as well with unreliable results. [9 - 10].

Feature selection can choose the most statistically significant attributes, which helps to build better models and to avoid overfitting. Hyperparameters are preset in ML models and can be tuned to fit specific situations and make better predictions. Combining these two methods enables models to make good forecasts that are cost- and resource-sensitive, reliable and actionable in medical fields as evidenced by Gao et al. [11].

Our primary research developed inventive feature selection and data processing techniques to improve predictions. We conducted systematic imputation strategies on a distribution of factors and used univariate analyses based on VIF and Random Forest methods. This resulted in better AUC-ROC than found in the literature. This study conformed with the TRIPOD guidelines, namely, Moons et al. [12] and Amritphale et al. [13]

## 2 METHOD

### 2.1 Data Availability

The MIMIC-III (version 1.4) database is an extensive, publicly accessible database that recorded 38,597 adult patients and 49,785 hospital admissions who stayed in ICUs of the Beth Israel Deaconess Medical Center in Boston, Massachusetts from 2001 to 2012. This dataset includes information on admissions, patient demographics, vital sign measurements, laboratory test results, procedures, medications, caregiver notes, imaging reports, and mortality (including dates and times).[14] The dataset is also deidentified therefore secondary which does not require approval by an institutional review board nor informed consent. [15])

### 2.2 Patient Selection

We restricted our study to 1,177 adult patients in the MIMIC-III database diagnosed with heart failure and who were admitted to the ICU. The target group initially consisted of 13,389 patients above 18 years of age selected using relevant International Classification of Diseases-9th Revision (ICD-9) codes. 162 patients without ICU admission were excluded. Another 4,871 patients, who did not have the N-terminal pro b-type natriuretic peptide (NT-proBNP) record were excluded from our study data, as NT-proBNP is a critical marker of heart failure. [16] Lastly, 7,179 were excluded for lack of echocardiography records, a basic tool for heart failure evaluation. [17] The process is detailes in Figure 1.

**Figure 1:**
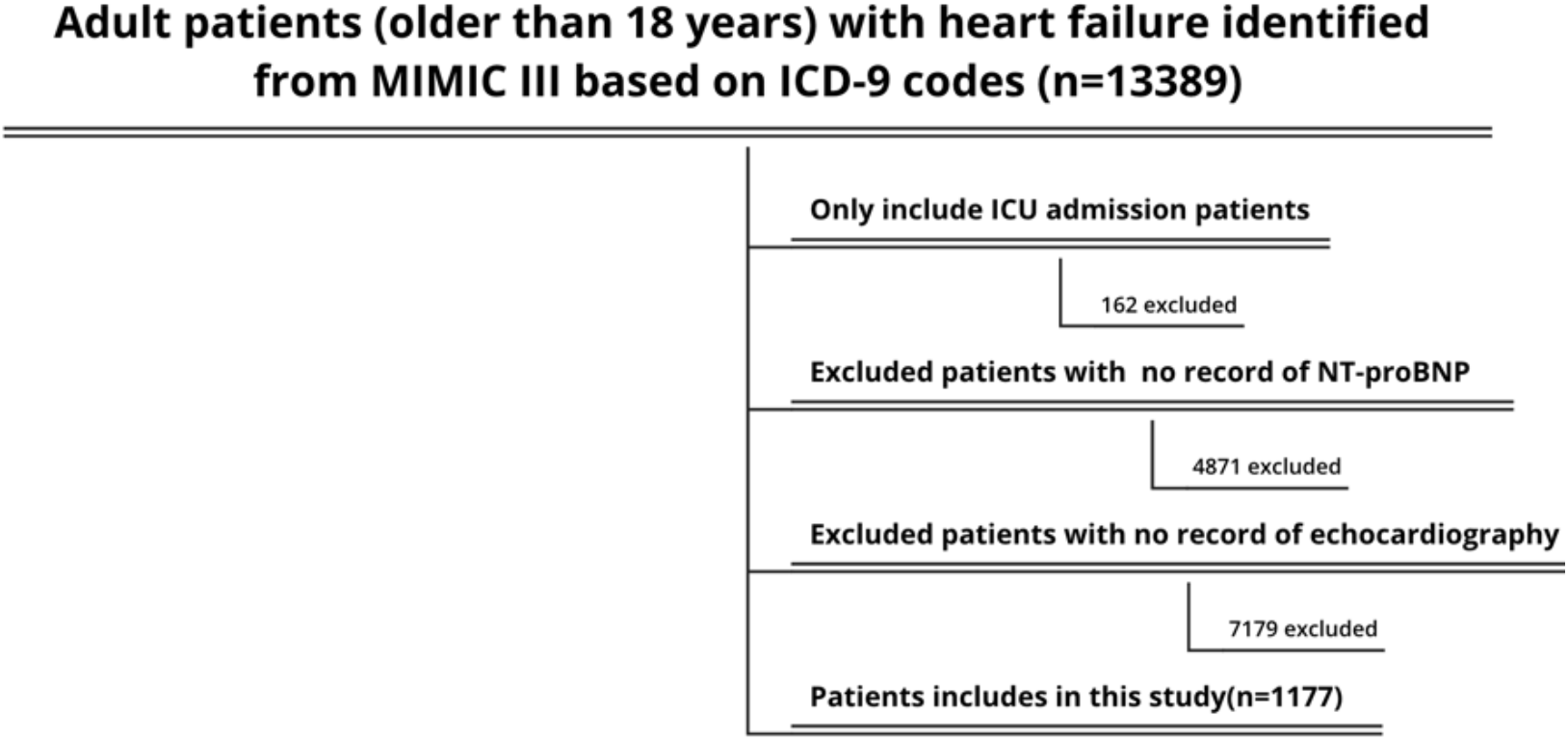
Flow chart illustrating data extraction process for study patient.

### 2.3 Feature Extraction

Using Structured Query Language (SQL) with PostgreSQL (V.9.6), demographic characteristics, vital signs, and laboratory values were extracted from the MIMIC-III dataset. Based on expert opinions, previous studies (Ashrafi et al. [18], Abraham et al. [19], Peterson et al. [20], Jia et al. [21], Lagu et al. [22] and Wang et al. [23]), and clinical relevance, 42 features were extracted from the original MIMIC-III database. Demographic characteristics and vital signs were recorded during the first 24 hours of each admission. Laboratory variables were measured throughout the entire ICU stay. Mean values were analyzed for features with multiple measurements. The primary outcome was in-hospital mortality, defined as 1 if the patient died during their ICU stay or 0 if the patient survived.

### 2.4 Pre-processing

We began the preprocessing stage by reading and cleaning the raw data obtained from the MIMIC-III database. We excluded missing values, columns with single unique values, and duplicate entries. Rows with null values in the outcome column were also discarded, and variables ‘group’ and ‘ID’ were deemed irrelevant.

For imputation, we used median imputation due to skewness and outliers in most features. We addressed outliers by selectively removing extreme data points. We then assessed the distribution of different classes within the outcome variable and identified an imbalance. To address this, we employed oversampling to balance the classes in the training set.

### 2.5 Feature Selection

The Variance Inflation Factor (VIF) was calculated to prevent multicollinearity in continuous features that could cause high standard errors in the prediction model (Murad et al. [24] and Lafi et al. [25]). Setting a lower VIF limit at 5, as suggested by Zach [26], we deleted variables with multicollinearity to reduce potential variables for modelling to 42.

LASSO regression was used for feature selection for numeric variables because it performaned best eliminating redundant or less informative features as detailed by Muthukrishnan et al. [27]. This process resulted in a small subset of numeric variables with importance scores greater than 0. We applied Random Forest for categorical data, as it best predicted accuracy for categorical data, and identified 10 categorical features with non-zero feature importance values. [28].

The univariate method based on LightGBM was used to order the significance level of all features. A threshold of 0.05 was used to eliminate features that contributed less, based on Bolón-Canedo et al. [29] This reduced the number of variables from 42 to 18 (see Table 1.)

**Table 1:** Summary of the features selected by category.

| Category | Feature |
| --- | --- |
| Demographic | BMI |
| Vital signs | Urine_output, Cystolic_blood_pressure, Temperature |
| Comorbidities | Atrial_fibrillation, Renal_failure, Depression, Hyperlipidemia, Deficiency_anemias, Hypertensive, COPD |
| Laboratory variables | Leucocyte, RDW, Basophils, Platelets, NT-proBNP, Magnesium_ion, Creatinine |
| Outcome variable | Mortality |

### 2.6 Modeling

We deliberately used a suite of machine learning models: Logistic Regression, LASSO Logistic Regression, Random Forest, LightGBM, Support Vector Machine, and Decision Tree, as they best analyze complex health data. Grid-Search was used to find the optimum set of hyperparameters for each model. Model performance was assessed by the AUC-ROC values against the test set.

Evaluation criteria included bootstrapped 95% confidence intervals of accuracy and AUC-ROC. Higher AUC-ROCs indicate better discrimination and confidence intervals to estimate model performances. These models facilitated exploring data resources and their elucidation. The Logistic Regression model, with the highest AUC-ROC and narrowest confidence interval, was proposed to predict ICU mortality in heart failure patients along with the other baseline models. The whole process is summarized in Figure 2.

**Figure 2:**
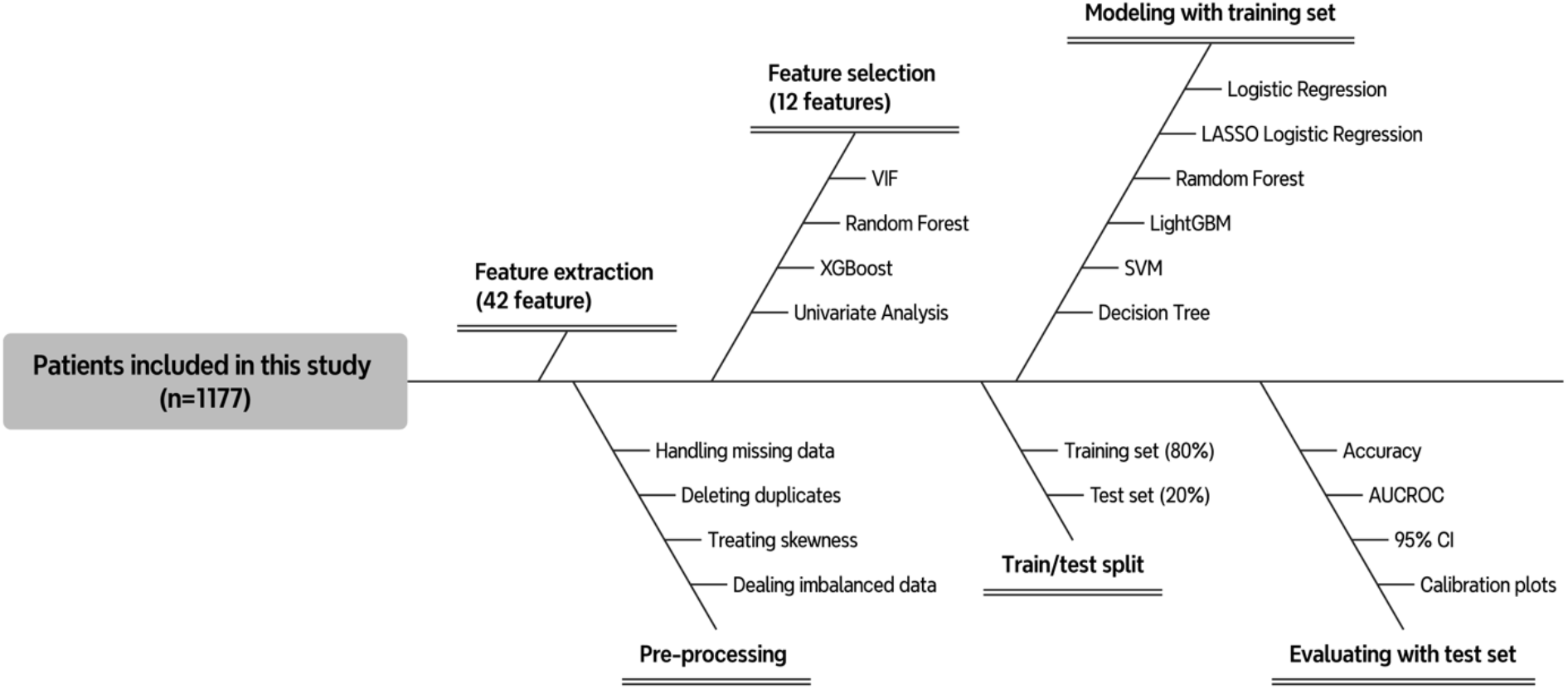
Flow chart illustrating study design.

## 3 RESULT

### 3.1 Model Evaluation

Table 2 summarizes results of our proposed model and baseline ML models using our evaluation metric. For our Logistic Regression model, the accuracy score is 0.7291 with AUC-ROC values of 0.7710 (95% CI 0.7458 - 0.7927) and 0.8766 (95% CI 0.8065 - 0.9429) for training and test sets respectively. Among baseline models, the LASSO Logistic Regression model performed best, yielding an accuracy score of 0.7291 and AUC-ROC of 0.7712 (95% CI 0.7462 - 0.7927) and 0.8754 (95% CI 0.8038 - 0.9420) in training and test sets (Figure 2).

**Table 2:**
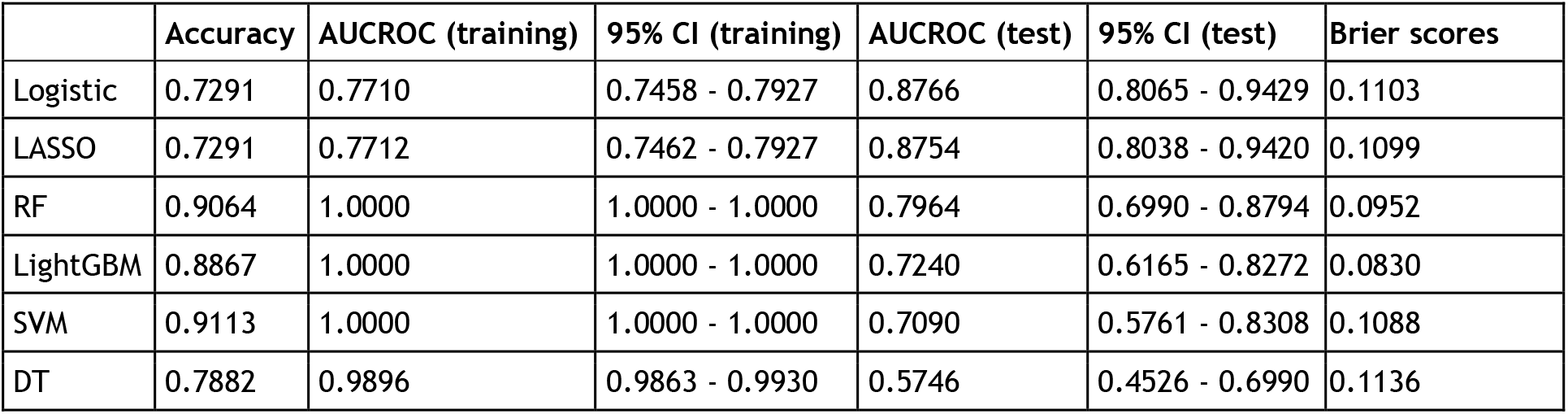
Evaluation results of proposed and baseline models.

|  | Accuracy | AUCROC (training) | 95% CI (training) | AUCROC (test) | 95% CI (test) | Brier scores |
| --- | --- | --- | --- | --- | --- | --- |
| Logistic | 0.7291 | 0.7710 | 0.7458 - 0.7927 | 0.8766 | 0.8065 - 0.9429 | 0.1103 |
| LASSO | 0.7291 | 0.7712 | 0.7462 - 0.7927 | 0.8754 | 0.8038 - 0.9420 | 0.1099 |
| RF | 0.9064 | 1.0000 | 1.0000 - 1.0000 | 0.7964 | 0.6990 - 0.8794 | 0.0952 |
| LightGBM | 0.8867 | 1.0000 | 1.0000 - 1.0000 | 0.7240 | 0.6165 - 0.8272 | 0.0830 |
| SVM | 0.9113 | 1.0000 | 1.0000 - 1.0000 | 0.7090 | 0.5761 - 0.8308 | 0.1088 |
| DT | 0.7882 | 0.9896 | 0.9863 - 0.9930 | 0.5746 | 0.4526 - 0.6990 | 0.1136 |

Calibration plots (Figure 3) and Brier scores (Table 2) highlighted the reliability of the predicted probabilities. The Logistic Regression model showed good calibration and a Brier score of 0.1103, indicating reliable predictions. The LASSO Logistic Regression and SVM models also had good calibration with Brier scores of 0.1099 and 0.1088, respectively. The Random Forest and LightGBM models had lower Brier scores (0.0952 and 0.0830). It shows these models are well-calibrated. The Decision Tree model, which had the highest Brier score (0.1136) and calibration, demonstrated reasonable predictive capability, albeit not as good as the other models. These results highlighted the importance of both AUC-ROC and calibration in evaluating model performance for clinical decision-making.

**Figure 3:**
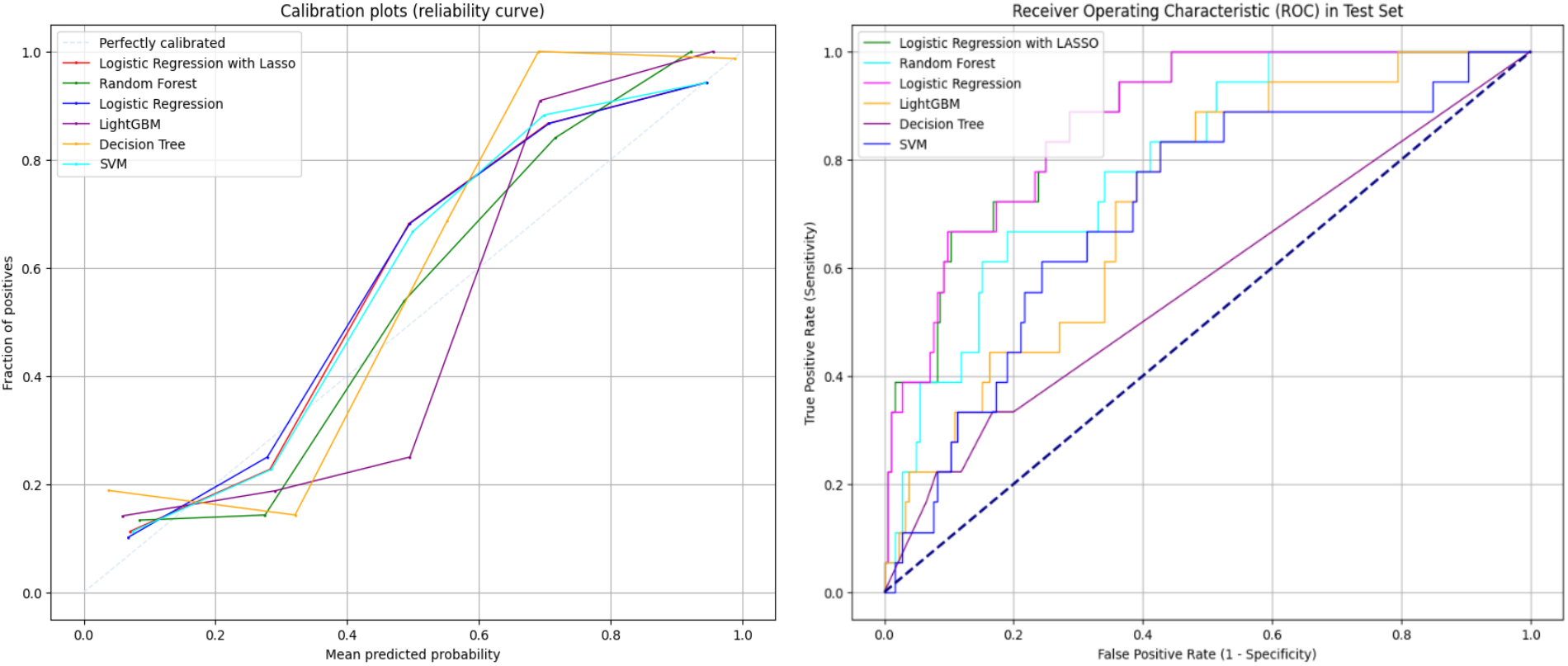
Receiver Operating Characteristic (ROC) Curve and Calibration Plots in Test Set.

### 3.2 Model Comparison

Logistic Regression and LASSO Logistic Regression models showed superior predictive performance. Based on Fein et al. [30], we used a within–subject t-test to determine the difference between the two models using the same dataset. We used 500 bootstrapped AUCROC score for the test. The null hypothesis was set to be “there is no difference between the Logistic Regression model and LASSO Logistic Regression model,” and the alternative hypothesis to be “there exists a difference between these two models.”. The small p-value of 1.0822×10^−43^ (Table 3) indicated we could reject the null hypothesis with 0.1% statistically significant level, meaning the models were dissimilar. The positive mean difference indicated that the Logistic Regression model had better prediction on average compared with LASSO Logistic Regression. Hence, among all the models we built, the Logistic Regression model was chosen for our proposed model with an AUCROC of 0.8766 (95% CI 0.8065 - 0.9429).

**Table 3.** T-test results using 500 bootstrapped AUCROC between Logistic Regression and LASSO Logistic Regression models.

|  | P-Value | T-Statistic | Mean Difference |
| --- | --- | --- | --- |
| Logistic vs LASSO | $1.0822 \times 10^{-43}$ | 15.3184 | 0.0013 |

### 3.3 SHapley Additive ExPlanations (SHAP) Analysis

SHAP analysis helped identify which features most influenced the ML model prediction, as suggested by Hamilton et al. [31]. Importance values were computed and presented by descending order (Figure 4). Leukocyte seemed to be the most crucial feature in mortality prediction of HF patients in ICU. The majority of points in red for Leucocyte, RDW, Creatinine, Magnesium_ion, NTproBNP, and temperature were positioned on the right side of the zero-center line. This suggests fatal outcomes for patients with higher values of these features. In contrast, parameters like Urinary output, Platelets, Basophils, BMI, and systolic blood pressure had more red points on the left side. This indicated ICU-HF patients exhibiting low values of these parameters had higher possibility of death.

**Figure 4:**
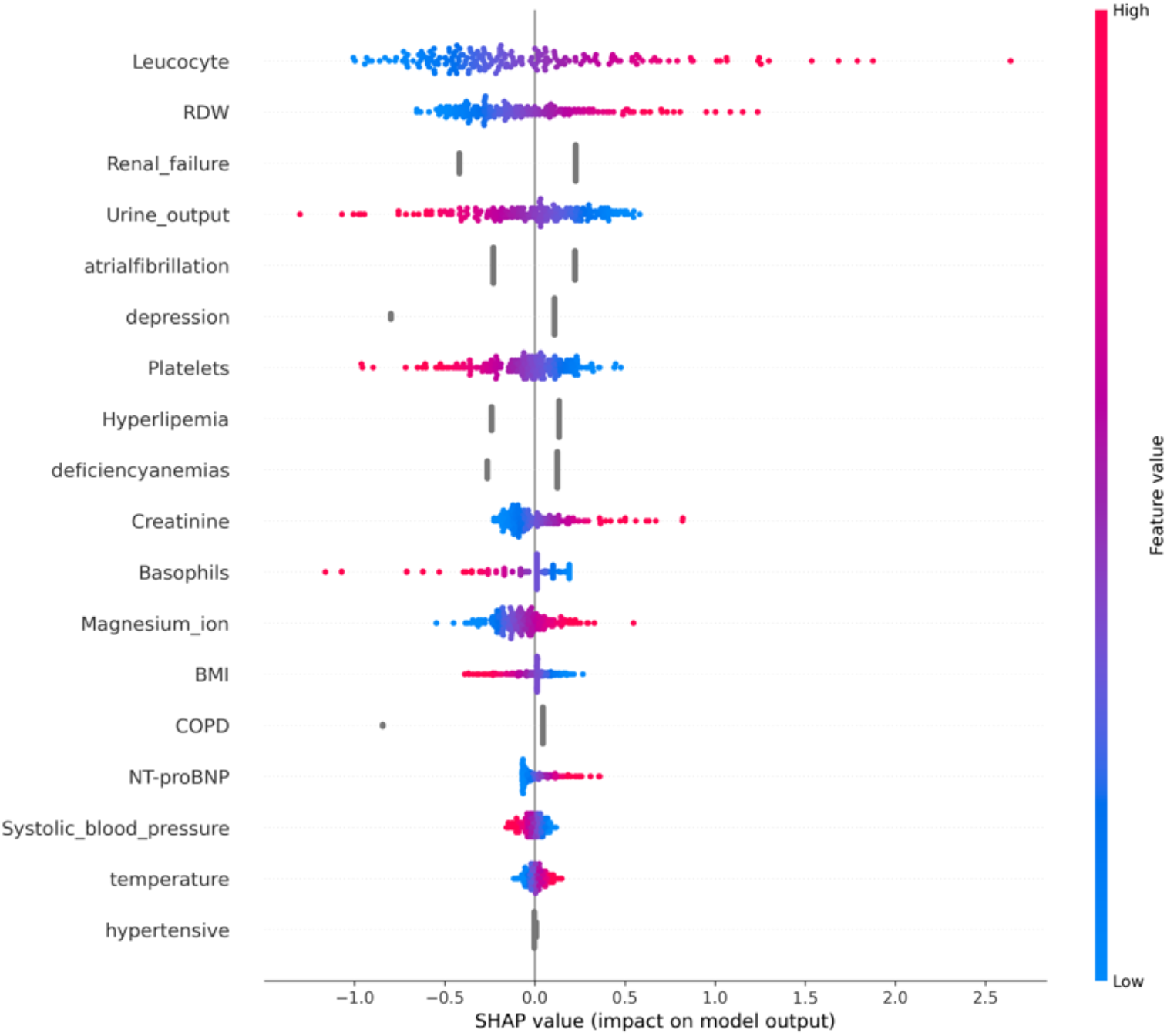
SHAP values for each variable using the proposed Logistic Regression model.

## 4 DISCUSSION

Many studies have tried to predict in-hospital mortality among ICU patients with heart failure using the MIMIC-III database. The majority of these studies though were limited by feature selection and imbalanced datasets. For example, Chiu et al. [32] utilized an ensemble algorithm to generalize a combination of models but had difficulties dealing with feature selection and managing imbalanced data.

Our study mitigated such issues using an innovative method of missing value imputation by equal distribution for one variable after a systematic imputation based on the median value of each variable. This minimized undesired effects of skewed data and outliers. Also, this study employed thorough feature selection techniques of VIF, LASSO, and Random Forest, and univariate analyses within the LightGBM framework. This method provided the most relevant features to improve the predictive ability of the model. Furthermore, the use of Grid-Search for hyperparameter fine-tuning guaranteed that each hyperparameter was set at its best level. The adjustments led to the impressive performance of the Logistic Regression model. Applying SHAP helped us to understand the decision-making process of the model, thereby promoting understanding of the clinical issues. The proposed Logistic Regression model achieved an AUC-ROC of 0.8766, a 9.18% improvement over the best AUC-ROC of 0.8029 reported by Li et al., who used XGBoost and LASSO regression models. [3]

## 5 CONCLUSION

This research focused on developing an ML model to predict the mortality of ICU patients with HF using data from the MIMIC-III database. We compared five baseline models to our proposed Logistic Regression model. The Logistic Regression model demonstrated superior performance over baseline models and the best existing models, achieving a higher AUC-ROC and a narrower 95% confidence interval. All models showed strong calibration and low Brier scores to validate their robustness and accuracy in predicting patient survival outcomes.

These enhancements were attributed to a rigorous feature selection process that reduced the initial set of features to 18 key variables. Comprehensive hyperparameter tuning using Grid-Search optimization ensured the best possible performance of the Logistic Regression model. SHAP analysis confirmed the clinical relevance of selected features, such as leucocyte count and RDW, further validating the model’s robustness.

Our framework offers valuable support to medical professionals by helping them identify ICU HF patients at high mortality risk. The predictive model evaluates the risk of death using various biomarkers, enabling clinicians to implement preventive actions effectively. This capability is particularly advantageous in critical care settings, where timely and accurate predictions can significantly impact patient outcomes.

## Data Availability

All data produced are available online

## Notes

### Competing Interest Statement

The authors have declared no competing interest.

### Funding Statement

This study did not receive any funding

